# Health and TB systems resilience and pandemic preparedness: Insights from a cross-country analysis of data from policymakers in India, Indonesia, and Nigeria

**DOI:** 10.1101/2024.05.09.24307131

**Authors:** Laura Jane Brubacher, Vijayashree Yellappa, Bony Wiem Lestari, Petra Heitkamp, Nathaly Aguilera Vasquez, Angelina Sassi, Bolanle Olusola-Faleye, Poshan Thapa, Joel Shyam Klinton, Surbhi Sheokand, Madhukar Pai, Charity Oga-Omenka

**Affiliations:** School of Public Health Sciences, University of Waterloo, Canada; TB PPM Learning Network, McGill International TB Centre, McGill University, Canada; Research Center for Care and Control of Infectious Disease, Universitas Padjadjaran, Bandung, Indonesia; Department of Public Health, Faculty of Medicine, Universitas Padjadjaran, Bandung, Indonesia; Research Institute of the McGill University Health Centre, McGill University, Montreal, Canada; Sustaining Health Outcomes through the Private Sector (SHOPS) Plus/Abt Associates, Lagos, Nigeria

## Abstract

**Introduction:** The COVID-19 pandemic was an unprecedented challenge to health systems worldwide and had a severe impact on tuberculosis (TB) case notifications and service delivery. India, Indonesia, and Nigeria are high TB-burden countries where the majority of initial care-seeking happens in the private health sector. The objectives of this study were to (1) explore policymakers’ perspectives on the impact of the COVID-19 pandemic on private sector TB service delivery in India, Indonesia, and Nigeria; and (2) identify cross-cutting lessons learned for pandemic preparedness with respect to TB service delivery.

**Methods:** From May – November 2021, thirty-three interviews were conducted with key policymakers involved in health service administration, TB service delivery, and/or the COVID-19 response in India, Indonesia, and Nigeria (n = 11 in each country). Interviews focused on the impact of COVID-19 on TB services and lessons learned for pandemic preparedness with respect to TB. Data were analyzed thematically using a hybrid inductive-deductive approach, informed by Haldane et al.’s (2021) Determinants of Health Systems Resilience Framework.

**Results:** Policymakers highlighted the crucial role of intersectoral collaboration, effective governance, innovative financing strategies, health workforce reallocation, and technological advancements such as virtual consultations and mHealth in strengthening TB service delivery amid the COVID-19 pandemic. India relied on patient-provider support agencies to implement a joint strategy for TB care across sectors and states. Indonesia engaged networks of private provider professional associations to facilitate coordination of the COVID-19 response. Nigeria implemented a pandemic policy for public-private referral for the continuity of TB care.

**Conclusion:** Countries implemented varied measures to support TB service delivery during the COVID-19 pandemic. This study presents lessons learned from three countries (India, Indonesia, and Nigeria) that together offer a ‘menu’ of possibilities for supporting pandemic preparedness with respect to TB care vis-à-vis strengthening health systems resilience.

## Introduction

COVID-19 and tuberculosis (TB) are the two deadliest infectious diseases globally, with 6.7 million and 4.2 million deaths from COVID-19 and TB between 2020 and 2022, respectively (1,2). Both are respiratory tract infections with some similarities in transmission, symptoms, and public health strategies (3,4). Since the pandemic, public health responses to TB have been severely impacted globally, as countries prioritized their COVID-19 responses over other public health functions (1,3).

The World Health Organization (WHO) Global TB Reports since 2020 highlight the complex impact of the COVID-19 pandemic on TB (5,6) and TB care (1). TB case notifications dropped globally in 2021, from 7.1 to 5.8 million cases between 2019 and 2020, with some increase to 6.4 in 2021 and a full recovery to 7.5 million in 2022. These drops were observed in most high-burden countries like India, Indonesia, and the Philippines, with a few countries in the African region showing some increases in notifications between 2019 and 2022. These reports indicate significant challenges in accessing TB care during the pandemic, with less impact in a few countries. India, Indonesia, and Nigeria are among high burden TB countries, with the highest proportions of initial care-seeking taking place in the private sector and with different trends in TB notifications since the pandemic (1). India and Indonesia experienced the highest and second-highest decreases in TB notifications due to COVID-19, respectively, while Nigeria reported increased notifications. This indicates some differences in TB case finding and pandemic responses within each country.

Diverse COVID-19 control measures implemented by different countries might have differentially impacted TB care. For example, countries like Nigeria that experienced previous outbreaks of infectious diseases like Ebola leveraged these experiences to inform their pandemic responses (7– 9). Studies have focused on the impact of COVID-19 on private sector care, specifically the varied responses and adaptations by providers (10–12). Indeed, while COVID-19 impacted TB services globally through disruptions to private healthcare provision and reduced patient turnout, the private healthcare sector also showed resilience and adaptability in continuing essential TB services despite challenges (13). The private sector had a crucial role in TB control efforts during the pandemic, as providers promptly adjusted services, implemented infection control measures, and maintained TB care quality (14).

Overall, countries adapted differently to the health system challenges presented through the COVID-19 pandemic; however, there is an opportunity to identify lessons from both the similarities and differences in how countries adapted, with an eye to strengthening ongoing private-public collaboration for TB service delivery for future pandemics. In 2023, United Nations High Level Meetings on TB (15), Universal Health Coverage (UHC), and pandemic preparedness emphasized the importance of building resilient integrated health systems. WHO is actively positioning TB care within a multisectoral framework (16,17) and emphasizes the importance of private sector engagement in UHC (18) as well as the Pandemic Accord and related governance systems (19).

The objectives of this study were to (1) explore policymakers’ perspectives on the impact of the COVID-19 pandemic on private sector TB service delivery in India, Indonesia, and Nigeria; and (2) identify cross-cutting lessons learned for pandemic preparedness and public health planning with respect to TB service delivery, using Haldane et al.’s (2021) Determinants of Health Systems Resilience Framework.

## Methods

### Study context

This qualitative study is part of a broader project, the COVID-19 Effect on TB in the Private Sector (COVET) study, which aimed to assess the post-COVID-19 landscape of private healthcare sectors in high TB-burden countries that have extensive private sector delivery of TB services: India, Indonesia, and Nigeria. The COVET study is a joint research project conducted by the McGill International TB Centre, Georgetown University, the University of Waterloo, Universitas Padjadjaran, and the USAID-funded Sustaining Health Outcomes through the Private Sector (SHOPS) Plus program. In India, the Institute of Socio-Economic Research on Development and Democracy and its partners leveraged the Patient Provider Support Agency (PPSA) network for data collection^1^. In Indonesia, the Research Center for Care and Control of Infectious Disease (RC3ID)-UNPAD collaborated with the local health office to utilize the study area of the 2019 Investigation of Services Delivered for TB by External Care Systems – Especially the Private Sector (INSTEP) study in Bandung (21). In Nigeria, the SHOPS Plus program facilitated partnerships for the study in Kano and Lagos (22). This broader project provided a robust methodological context to comprehensively examine the post-COVID-19 private sector TB landscape across diverse settings.

### Data collection

From May to November 2021, key informant interviews were conducted with policymakers involved in private sector TB care in India, Indonesia, and Nigeria, recruited through COVET study networks and partnerships (n = 33 policymakers; n = 11 interviews per country). Interview participants were sampled purposively according to their roles and responsibilities within government TB programs, communicable disease programs, general health service administration, private sector professional associations, and NGO program administration and selected as a result of a stakeholder mapping exercise conducted by the research team in each country (Table 1). Interviews included policymakers’ perspectives on the impact of the COVID-19 pandemic on TB health service delivery, the performance of their pandemic responses, and lessons learned from the impact of COVID-19 on TB services in their respective countries (see Appendix A for a sample interview guide). Interviews were conducted virtually in adherence to COVID-19 safety guidelines and in English, except for one interview each in Hindi (India) and Bahasa (Indonesia), as per the participants’ preferences.

**Table 1.**
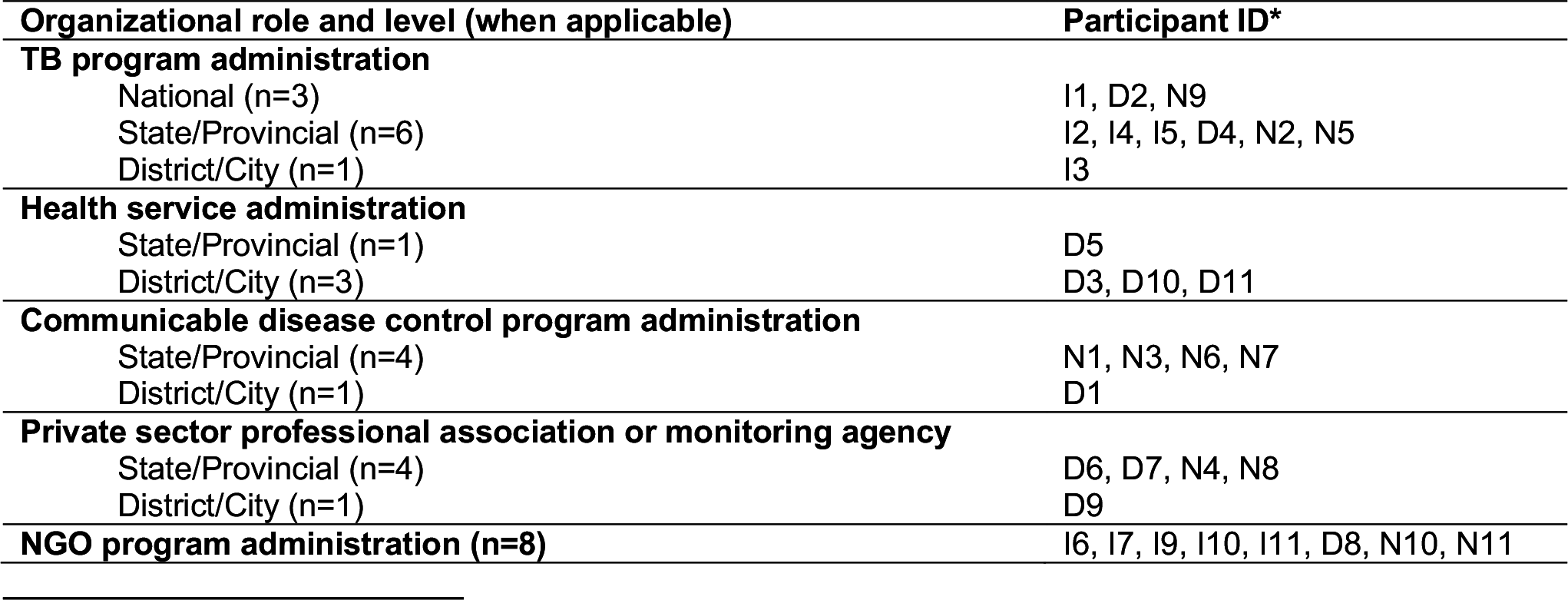
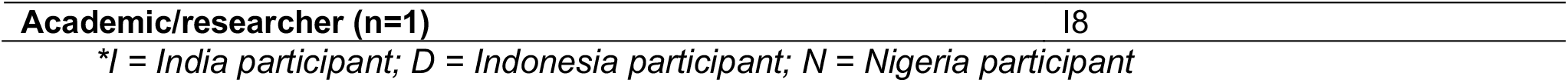
Key informant interview participants’ organizational roles or affiliations.

Interviews in Hindi and Bahasa were subsequently translated and transcribed into English. Interviews were approximately 50 minutes in duration (range: 30-60 minutes) and were audio-recorded with permission. All individuals provided verbal informed consent to participate. Ethics approval was obtained from McGill University (Certificate #: COVID BMGF/ 2021-7197), Georgetown University (STUDY00003422), Universitas Padjadjaran (No.166/ UN6.KEP/EC/2021), HREC Lagos State University Teaching Hospital (LREC/06/10/1517), and HREC Kano State MoH (MOH/Off797/T.I/2168). Patients or the public were not involved in the design, conduct, reporting, or dissemination plans of this research.

### Data analysis

Data were analyzed thematically, using a constant comparative approach within and across interview transcripts, to identify insights into COVID-19 impacts and adaptations (23). A hybrid inductive-deductive coding approach was employed (24). Research teams in each country conducted an initial inductive thematic analysis. Subsequently, teams met to compare and contrast their coding and develop analytic insights across countries (25). This analysis created the foundation for subsequent secondary analysis by the University of Waterloo team to map inductive codes to Haldane et al.’s determinants of health systems resilience framework (20). The use of this framework allowed us to identify cross-cutting lessons learned for strengthening health system preparedness for future pandemics with respect to TB services. *NVivo®* software was used for organizing and retrieving codes and coded excerpts. Collaboration among the research team and peer debriefing contributed to the validity of the analysis (26).

## Results

Policymakers’ perspectives on the impact of the COVID-19 pandemic on private TB services and lessons learned mapped to the following domains of the health systems resilience framework (20): the cross-cutting domain of intersectoral collaboration (which we have adapted to: increased public-private health sector engagement); governance and financing; health workforce and health service delivery; public health functions; and medical products and technologies.

### Increased Public-Private Health Sector Engagement

Across countries, policymakers underscored how COVID-19 amplified public-private sector engagement, with implications for TB care moving forward. In Indonesia, a participant from the private sector noted the reliance of the public sector on private clinics to ‘roll out’ the COVID-19 vaccination program – that it was a *“blessing in disguise”* that the public sector had to determine how to utilize and collaborate with private clinics: *“In the beginning, they heed[ed] no attention towards clinics, but eventually they asked for our help. And this whole thing changes the paradigm – after all, we are present not as competitors”* (D6). Similarly, Nigerian policymakers reported an “all hands-on deck” public-private health sector collaboration evident in the pandemic that can be leveraged into TB service delivery (N9). This built upon a strong pre-existing role of the private sector in TB care, which provided a ‘cushion’ to fill gaps when public sector care was difficult to access during pandemic lockdowns (N5). In India, PPSAs actively collaborated during the pandemic with the National TB Elimination Program (NTEP) to develop and implement a joint strategy for TB service delivery across States and sectors (I1). Beyond case notifications, one policymaker in India also described how COVID-19 opened and improved existing lines of communication between sectors (vis-à-vis a Deputy Commissioner’s communication with the private sector regarding district COVID-19 transmission), which may support TB service delivery in the future (I3). Based on the experience of private-public health sector collaboration in COVID-19, a policymaker in India also described what an increased public private mix could look like moving forward, with relevance to TB service delivery:

Ultimately, the heads of the organizations either in the public or private sector, there will be a face – whoever it is. For example, it’s the director of health services and there would be a similar representative from the private sector. I think that level of conversation of the local IMA [Indian Medical Association] with government was very critical for information to percolate. Unless the top people are convinced, it would not go down. I think those meetings with heads of organizations and then percolating it and cascading it down to their respective members and units was probably the best way (I7).

Overall, there was evidence of the crucial role played by the private sector in TB care during COVID-19 and an identified need for continued strengthening of public-private health sector engagement for the purpose of: (a) augmenting the public system and filling gaps in TB service delivery through the private sector vis-à-vis reallocation of the health workforce, extending the ‘reach’ of care, and extending the capacity of the health system overall; (b) dissemination of guidelines, policy, and training for consistency and clarity of approaches to TB care amid a pandemic, across public-private health sectors; and (c) expanded distribution networks for personal protective equipment (PPE), TB medications, and vaccines through private facilities (described in detail below). This importance of private health sector engagement in TB care, as illustrated in the COVID-19 pandemic, is integrated into the following sections – which map to the core domains of the determinants of health systems resilience framework.

### Governance and Financing

The crucial role of private provider professional associations or NGOs as intermediaries between the public sector and a network of private facilities (i.e., PPSAs in India) was highlighted across all three countries. In Indonesia, standardizing COVID-19 protocols (i.e., PPE usage) and overall coordination and communication across facilities occurred via associations like ASKLIN (private clinic association). While these associations were seen as ‘nodes’ through which increasing public-private engagement was observed in COVID-19 (D6), respondents also felt the public sector predominantly sets standards for private clinics. For instance, PPE requirements were set without any government support for private providers, which increased their operational expenses (D5). A policymaker noted the need to incentivize the private sector (through finances, infrastructure, and tools) to implement government programs, highlighted in the pandemic and relevant to TB services (D4). Similarly, in Nigeria, the networks of professional associations were leveraged to disseminate policies and practice guidelines from the public sector and distribute PPE. A central government agency (Health Facility Monitoring and Accreditation Agency, HEFAMAA), which provides performance monitoring/accreditation to private facilities, was also leveraged to disseminate COVID-19 infection prevention and control protocols through its private provider networks. Nigeria also established a more formalized structure for integrated TB and COVID-19 response across public and private sectors vis-à-vis a “State TB-COVID-19 response team” (Kano State). In India, policymakers identified a need for more foresight as to how to leverage the resources and capacity of the private sector as well as a general need for enhanced partnership and lines of communication across sectors rather than unilateral communication (i.e., from the NTEP to private providers). This was underscored in the COVID-19 pandemic but has relevance to TB.

Overall, as stated by a policymaker in India, there was a political commitment to a multi-sectoral, community-engaged, and collaborative approach to the COVID-19 pandemic response, which could be leveraged and replicated to support progress in TB service delivery:

Things will not happen in the field just by issuing letters and guidelines and unless we sit with the states and be a part of their implementation, it is not going to happen. The technical support agencies, the PPSAs, the civil society partners, the community partners, a lot of them, all of them joined hands to give a response to COVID. And if we can do that, all of government approach for COVID, then we can do that for TB as well. That is what we are going forward with. And there is absolutely full commitment from the administrative setup, from the political setup and from the implementers. All the three are trying to go in one direction (I1)

### Health Workforce and Health Service Delivery

Policymakers across countries identified the need for reallocation of health human resources amid the pandemic, to cover service gaps due to staff shortages and ensure continuity of care for other diseases, like TB. A Nigerian policymaker observed that this *“task shifting”* of healthcare workers occurred more readily within the private sector than the public whereby *“sometimes there’s a vacuum or disruption in service”*, as was observed during the pandemic when health human resources were directed towards COVID-19 prevention and treatment (N11). In Indonesia, a centralized system of mapping and distributing the health workforce across the public system existed, whereby there was coordination between public health facilities, especially at the primary care level like *Puskesmas (Pusat Kesehatan Masyarakat)*, to cover gaps (D3). A policymaker in India identified how training public sector doctors and district TB officers, alongside creating awareness to the public about the availability of TB diagnosis in COVID-19 in government facilities, has increased case notifications (I4).

Countries employed a variety of strategies, engaging different organizational ‘actors’ to facilitate treatment continuity for TB patients amid COVID-19. In India, the NTEP implemented a large-scale telephone follow-up and TB treatment intervention to ensure door-to-door delivery of treatment (I4) as well as adherence support through call centres (I6). Health sector NGOs such as Alert India and PPSAs communicated with patients regarding the availability of TB medication and actively sourced public sector drugs for patients unable to access private sector drugs in COVID-19 (I5, I7). Nigeria had a policy that allowed referral of TB patients from the public sector to nearby private hospitals for continuity of care (N10); longer prescription refills given across private and public facilities to avoid unnecessary visits (N2); home-based care of patients (N6); and, overall, less of a *“dividing line”* between public and private sectors to ensure treatment continuity. A Tuberculosis and Leprosy Supervisor (TBLS) within the NTP followed up with TB patients to support treatment adherence irrespective of whether patients accessed care at private or public health facilities (N11). In Indonesia, private clinics often refer TB patients to public sector facilities (i.e., *Puskesmas*). Policymakers emphasized that while the pandemic increased overall communication between private and public sectors, it also underscored the need for enhanced reporting mechanisms and TB data synchronization to reduce patient loss to follow-up (D6, D8).

### Public Health Functions

Across countries, the pandemic enhanced the capacity of the health workforce to conduct contact tracing for COVID-19 and, consequently, active case finding for TB. One policymaker in India noted improved *“overall health system capacity to deal with communicable disease”* (i.e., through staff skills in outbreak management and resources) and that new molecular diagnostic technologies were purchased for COVID-19 which could be leveraged to support TB diagnostics (I1, I2). Similarly, in Nigeria, the pandemic reiterated the importance of active case finding and testing – for COVID-19 and TB – alongside increased public health awareness of these diseases’ similar symptomatology. To do so, Nigeria engaged multiple actors (i.e., SHOPS Plus) in community education and health promotion (N2).

The COVID-19 pandemic enhanced monitoring/reporting mechanisms, as shared by policymakers in India and Nigeria. In India, new mechanisms were established for virtual information exchange and reporting for TB, such as WhatsApp groups among private providers for addressing questions related to TB and COVID-19, as well as mobile applications designed for TB reporting by Patient Provider Support Agencies (PPSAs) (I10). Nigerian policymakers noted their already well-established monitoring and evaluation infrastructure (i.e., an integrated data surveillance system) within the TB program at the local government level and across the private and public sectors, which can continue to be leveraged for both TB and pandemic response. Also, in light of the pandemic, TB monitoring and evaluation staff now have the capacity to work virtually, which may confer longer-term benefits in terms of workplace capacity and efficiency (N3, N11).

### Medical Products and Technologies

Telemedicine technologies were implemented or more effectively utilized amid COVID-19 across countries (public sector platforms included *e-Sanjeevini* in India; *Halodoc* in Indonesia; and *EkoTelemed* in Nigeria)^2^. Private sector facilities developed their own mobile applications for virtual consultations; mobile applications were also used across sectors for support with TB prescription renewal and treatment continuity. In India, COVID-19 emphasized a need to invest in, mobilize, and scale-up information technology infrastructure to facilitate TB treatment adherence in a pandemic (e.g., through video-based directly observed treatment). This would require accompanying training for providers and patients in the use of these technologies. A policymaker suggested that teleconsultations could be implemented through PPSAs, which are already involved in ensuring TB treatment adherence (I5). Similarly, Indonesian policymakers identified a need for regulatory change regarding telemedicine or online consultations being recognized as ‘visits’ to facilitate funding to private facilities (D4).

Overall, each country shared evidence of intentional efforts and strategies for coordinating TB care within the COVID-19 pandemic, as well as opportunities for enhancing this coordination for future pandemic preparedness. For example, policymakers described how the public sector in India relied on private professional associations to implement training for private healthcare workers on TB and COVID-19 infection prevention and control measures (I1). Indonesia strengthened their TB lab network amid COVID-19 and disseminated a circular letter that regulated hospitalization of drug-resistant TB during the pandemic (D11). Nigeria created a State-level formal structure (within Kano State) for integration of the TB and COVID-19 response, referred to as the State TB-COVID-19 response team (N5). Policymakers across countries still also identified a need for more consistent and coordinated management of TB within a pandemic setting, in the form of a national-level protocol for TB management in a pandemic (N9); enhanced public-private regulation for TB management (D11); or more intentional leveraging of resources from a substantive private sector to support standardized COVID-19 protocols across public and private health facilities (I10). The lessons learned from COVID-19 and TB management that were identified by key policymakers across the three countries mapped closely to Haldane et al.’s (2021) health systems resilience framework (with health workforce and health service delivery considered together) (Figure 1).

**Figure 1.**
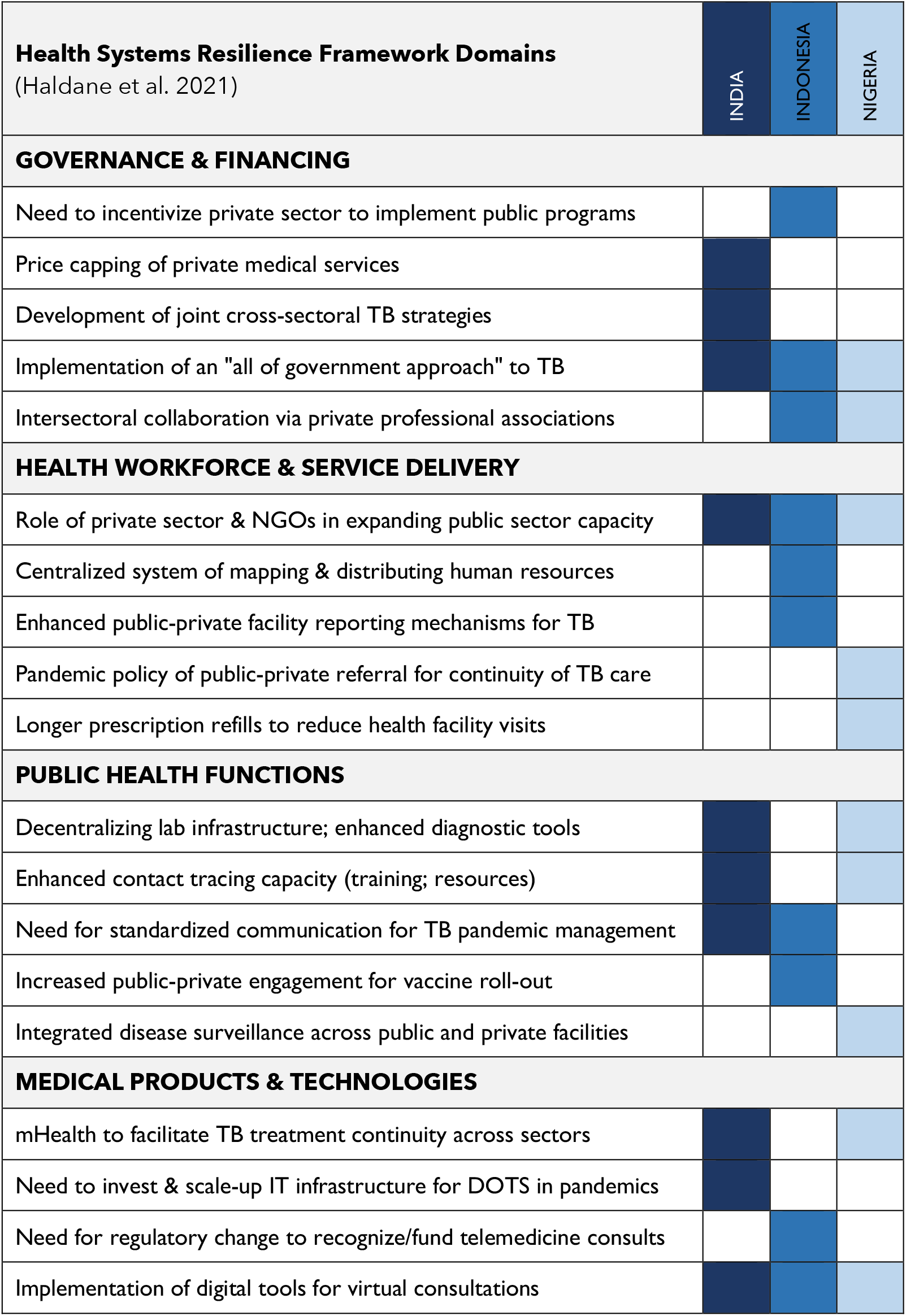
Lessons learned on health systems resilience and pandemic preparedness for TB care.

## Discussion

As our findings indicate, the private sector in India, Indonesia, and Nigeria galvanized in the COVID-19 pandemic to provide TB services. During the pandemic, when public health systems were in disarray, the private health sector mobilized to fill crucial gaps in TB service delivery and to augment the public sector workforce, as well as public health functions (i.e., testing, active case finding). Indeed, the strengthening of the private sector that occurred during the pandemic has contributed to current progress in public-private partnerships (14,27). These findings align with recent studies that similarly underscore the importance of the private sector in providing care for TB patients during a time of uncertainty in which public facilities were overwhelmed (10,25,28).

While our findings reveal differing pandemic control and preparedness strategies to varying degrees across countries, we found three major themes across the three countries. These include the implementation of an ‘all of the government approach’ to TB as part of governance and financing; the role of the private sector and NGOs in expanding public sector capacity, particularly for the health workforce and service delivery; and the implementation of digital tools for virtual consultations, as part of TB technologies. These findings align with studies from other countries, including high TB-burden countries. Studies have underscored the significance of a robust national strategy in combating the pandemic and how this shows promise for improved TB management and enhanced preparedness for future pandemics (20,29,30). The role of the private sector in maintaining services for TB during the pandemic and the increases – although short-lived in most settings – are well documented (5,11,14,31,32). Our findings also point to the mechanisms (or ‘nodes’) through which public-private health sector engagement for TB care was occurring during the COVID-19 pandemic. The public sector interfaced with private providers’ professional associations across study countries (i.e., ASKLIN, ARSSI, PERSI in Indonesia), as well as with PHIMA in Nigeria and PPSAs in India. Opportunities may exist to focus resources on these existing networks, leveraging these partnerships, to enhance TB services (13,14,33).

Several authors have synthesized global data on health systems resilience and pandemic preparedness during various epidemics, and highlighted the need for global coordination; guidance, collaboration, and information sharing; strengthening weak primary healthcare; acting on social determinants of health; and early, rapid, and aggressive actions in public health interventions (20,34–38). Similarly, the WHO Preparedness and Resilience for Emerging Threats (PRET) initiative calls for an integrated approach to pandemic response, health systems strengthening, enhancing national capacities for planning, coordination, risk communication, community engagement, health intelligence, and interventions, as well as global collaboration and expert consultations (39). In our study, various features of countries’ overarching governance approaches to the COVID-19 pandemic were highlighted as being relevant to TB delivery moving forward: most notably, the espousing of an ‘all of government approach’ to COVID-19 that could be leveraged into TB. This multi-sectoral, broadly cooperative approach, evident particularly in the early stages of the global COVID-19 pandemic response, has been recognized within the health systems resilience and pandemic preparedness literature as a governance element facilitative of improved outcomes (20,34,40,41).

Importantly, with respect to TB service delivery, there will not be one approach that fits every context. Rather, our study presents lessons learned from three countries (India, Indonesia, and Nigeria) – some were cross-cutting, and others were unique to one country’s context. Together, these findings offer a ‘menu’ of possibilities for supporting pandemic preparedness with respect to TB care vis-à-vis strengthening health systems resilience. However, the reality and practice of these findings are left to be seen, and TB systems have yet to fully leverage COVID-19 pandemic innovations (42). Working on cross-cutting and health systems related topics is not natural to disease programs and requires innovation, insights from other disciplines, and entrepreneurial initiatives. On top of a burnt-out health workforce, with decreasing funding and support, such a demand will be a substantial challenge.

### Strengths and Limitations

A strength of this study was that it examined lessons learned from the COVID-19 and TB response across three countries, as reported directly by key policymakers involved in these responses. Our study highlighted the importance of policymakers focusing on collaborative governance, innovative financing, workforce reallocation, and leveraging technology to enhance service delivery for pandemic preparedness, as well as the lessons from successful strategies to improve patient-provider support and private provider engagement. However, this did not include the perspectives of individuals and providers involved more ‘downstream’ in public health and primary care provision who may have offered distinct insights with respect to service delivery and pandemic adaptations or those more involved in public sector services. Further research is needed that retrospectively examines the COVID-19 response as pertaining to TB services from more diverse perspectives and analyzes the evolution of this response – and lessons learned – following this study’s data collection time point of 2021. Further research is also needed that examines the cost-effectiveness of infection control measures, analyzes TB staffing needs and constraints amid pandemic response, and explores the possibilities of strengthening lab networks to support both outbreak control and service delivery.

## Conclusion

In conclusion, this study provides a comprehensive analysis of the intricate interplay of public-private mixed health systems between the COVID-19 pandemic and tuberculosis (TB) care in three high-burden countries: India, Indonesia, and Nigeria. The WHO Global TB Reports for 2021 and 2022 underscore the challenges in accessing TB care during the pandemic, marked by initial global TB case notification reductions followed by subsequent increases (43,44). Notably, the three nations exhibited distinct trends in TB notifications, emphasizing the dynamic nature of pandemic responses within each context.

Reflecting the global UNHLM political declaration and the WHO guidance in the TB-MAF, our interviews with key policymakers highlighted the crucial role of intersectoral collaboration, effective governance, innovative financing strategies, health workforce reallocation, and technological advancements in strengthening TB service delivery amid pandemics. As we navigate the complex landscape of TB care, intersecting with the challenges posed by COVID-19, these lessons serve as a pertinent guide for policymakers, offering an array of possibilities to enhance health systems resilience and pandemic preparedness in the realm of TB services. This study underscores the importance of a collaborative, community-engaged approach as we strive for more effective and adaptable TB care in the face of future health crises. It will be key to continue documenting the implementation of such approaches to guide national health systems and TB programs with concrete and practical steps in achieving resilient systems that are prepared for future pandemics.

## Data Availability

All data produced in the present work are contained in the manuscript.

## Acknowledgements

Thank you to all staff of SHOPS Plus, the TB PPM Learning Network India, and Universitas Padjadjaran who supported data collection for this study.

## Footnotes

1 The PPSA is a health service delivery model in India that involves contracting a third-party agency (i.e., NGO) to engage private sector providers in TB service provision (45).

2 Maintenance or continuation of these technologies at the time of publication was not researched.

