## Appendix A for "Health and TB systems resilience and pandemic preparedness: Insights from a cross-country analysis of data from policymakers in India, Indonesia, and Nigeria"

**Appendix A:** Policymaker interview guide used and adapted slightly for data collection in each country: India, Indonesia, and Nigeria.

***Roles and responsibilities of key informant***

1. What is your job title, state or federal department/organization, and what city do you work in?
2. What type of facilities/operations do you oversee?
3. What are your main duties, especially with regards to TB in the private sector?

4. In what ways are you involved in policymaking and/or enforcement of policies for private health facilities and providers?

   *Optional follow-up/probing questions (especially if you're not getting a clear sense of the informant's role):*
   - Can you tell me about a typical day or week at work?

5. How would you describe you/your agency's level of interaction (communication) with private health facilities?

   *Follow-up/probing questions*
   - How often does your department meet with or communicate with the private sector providers?
   - How many private sector providers does your department communicate with (i.e., approximately how many providers do you meet with (or have met with)?

***COVID impact on the private sector***

6. Based on your observations, how has the COVID-19 pandemic impacted private healthcare providers' services in general?

   *Follow-up/probing questions*
   - Were practices shut/open/changed? Temporarily or permanently? Has this changed over the months? What percentage of facilities were affected? Have visiting hours for practices changed?

7. In what ways are these impacts/changes affecting people affected by TB in your jurisdiction?

   *Follow-up/probing questions*
   - Are providers offering the same services? Do private providers insist on a test for COVID-19 before testing for TB?

8. What new regulations/policies/procedures has your department issued in response to the COVID-19 pandemic that directly affect private facilities?

   *Follow-up/probing questions*
   - Types of policies (distancing, PPE, treating COVID-19 cases, price caps, purchasing services from private sector providers, other respiratory diseases)?
   - Timing of policies (e.g., still in effect? Only in effect during active lockdowns?)
   - Is there any policy in place that allows your department to take over any private facilities for COVID-19 care? If yes, what factors made you and your department decide to take over these facilities or decide not to take them over?
   - Is there any policy in place that allows the federal or state government to implement price caps to private sector providers? If yes, were these policies used during the

pandemic for COVID-19 or other health services besides COVID-19? If yes, what other health services did the price caps apply to?

9. How is your agency/department enforcing these regulations/policies?

10. Are there any other COVID-19-related legislations/policies/regulations not under your department/agency's purview that directly affect private health facilities?

***Private sector response and performance***

11. What has been the response from providers regarding these policies/regulations?

    *Follow-up/probing questions*
    - If there has been a negative response, how has your department addressed these concerns?

12. How satisfied are you with private sector engagement in COVID-19 so far?

    *Follow-up/probing questions*
    - *Do you have any concerns about the performance of the private health care sector in the light of COVID-19?*

13. What are you and your department anticipating as next steps in the coming months?

---
